# A connection between factors causing diseases and diseases frequencies. It’s application in finding disease causes

**DOI:** 10.1101/2023.04.30.23289320

**Authors:** Alan Olan

## Abstract

In this work an author is building a theoretical model of a non-infectious disease, which shows that there is a connection between *diseases frequencies and their causes*. This connection allows to determine how many factors are causing a specific non-infectious disease if we know the disease rate in a population. The model shows that for a majority of non-infectious diseases there are at least two simultaneously acting factors which cause a disease and in many cases there are more factors simultaneously involved. This helps researchers to improve an understanding of specific disease causation and physiological mechanisms behind it and will lead to a research for additional, still missing causes to complete these mechanisms. This work determines a number of simultaneously acting factors causing diseases such as a breast cancer, coronary heart disease(CHD), multiple sclerosis, etc. and explains so called French Paradox for CHD. The work also deduces a formula and a *method* of *determining* that a specific *risk factor is the one which really causes a disease* or it is not. Applying a method developed in this work the author shows how three different simultaneously acting causes of atrial fibrillation are determined using an existing research data. This method should allow medical researchers to determine if a found risk factor for a disease is really a *cause of the disease* or not and covers a significant gap in current understanding of risk factors nature and its connection to the physiological parameters of the human body..

**Summary:**

- Using statistical and experimental data a mathematical model of non-infectious disease is created which is applicable to any non-infectious disease (and to some degree to infectious diseases as well)
- The model is considering that **a cause of a disease is *a change* in a physiological parameter** of the body approximately beyond 1-sigma interval of its measurements, slightly less.
- The model predicts and the experimental data confirm its predictions that rate of non-infectious diseases is closely connected to the number of changes to physiological parameters of the human body which are causing the disease. Based on this connection **<u>the model allows to determine number of disease causes (as number of physiological parameters’ changed) for any non-infectious disease by only knowing the disease rate</u>**. For example, if the disease rate is 72 per 1000 people then the model determines that disease has 2 causes. *Despite the differences in the rates* of a specific disease in different countries or populations the model determines the *same* number of causes. The model introduces a formula to calculate a number of disease causes as below:

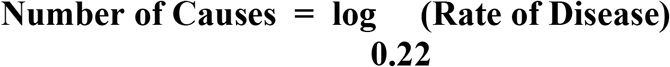

where a rate of disease can be for example as 45 per 10000 people and number of causes(as physiological parameters’ changes) is an integer number like 1, 2, 3, etc.

- According to this non-infectious disease model in order **to cause a disease *all* of specific for each disease physiological parameters should be changed by actions of the external environment so their measurements will be beyond 1-sigma interval**. Actually, slightly less than this interval.
- The model shows **<u>the more the rate of non-infectious disease the less number of simultaneous changes to the physiological parameters is required to cause it</u>**. The changes can be acquired over the lifetime of individual and have to be such so each physiological parameter’s measurement goes **beyond 1-sigma interval**\*.
- The work also allows to use vast volume of existing medical research’s **data about diseases’ risk factors in order to determine real causes of non-infectious diseases** using a simple criteria which *mathematically* derived from the model.
- The work mathematically **derives the criteria which allows to determine which risk factor is a cause of disease** and which is not. It mathematically proves that in order **<u>to be a cause of the specific disease the found risk factors</u>** (calculated as **[ risk_cases - normal_cases ] / normal_cases**) **<u>should rich the value of 3.5 (or 350%)</u>** or **19.7** (1970%), etc. plus/minus a practical margin (recommendation is +/-50%)
- The calculations based on the **<u>model has shown that a majority of non-infectious diseases are caused by at least 2 simultaneously occurring changes to physiological parameters of human body</u>** or more. For example it shows that **Coronary Heart Disease (CHD) is caused by 4 physiological parameters changes taking place at the same time**.
- As the model derived in this work predicts that non-infectious diseases are caused at least by 2 changes (beyond 1-sigma) to physiological parameters of human body then it shows there is no reason and actually, often it is incorrect to search for a single cause of the non-infectious disease. **The single cause of non-infectious disease does not exist for a majority of them**, according to this mathematical model.
- Based on the smoking risk factor value, the developed in this work **criteria determines that smoking is one of the causes of lung cancer** and that is matching to already a well recognized medical fact, and supports the validity of the criteria used to make this prediction.
- The model shows that Multiple Sclerosis in men is caused by changes to 5 physiological parameters (beyond 1 sigma interval *) while the same disease is caused by changes to 4 physiological parameters in women. This **explains why a rate of the Multiple Sclerosis is higher in women than in men** as the more physiological parameters needs to be changed to cause the disease the less the rate of disease.
- The work shows why **breast cancers** and **leukemias** are **caused by a change to 6 physiological parameters of the human body**.
- The model leads to a conclusion that the majority of non-infectious diseases cannot occur if only one physiological parameter of human body changes beyond 1-sigma interval. **The multiple and simultaneously taking place changes to physiological parameters needs to be there for a non-infectious disease to occur** but the changes can be acquired over the time.
- The work leads to a conclusion that by controlling few physiological parameters of the human body so they are located within the 1-sigma of its measurements **it is possible to prevent a non-infectious disease such cancers or strokes from occurring at all** in the individual.
- The work shows that the fact that few of physiological parameters changes needed to simultaneously occur in order to cause the non-infectious disease, **allows to select some risks factors as real causes of the disease** and that we have some room for an error in selecting these risk factors as disease causes because if few risk factors are correctly chosen they will compensate for an incorrectly chosen one. This way we can prevent many diseases from occurrence by keeping the right risk factors (physiological parameters) under control (meaning within 1-sigma interval).
- The work mathematically derives a **<u>formula which connecting the numbers of disease causes</u>** (physiological parameters’ changes) **<u>determined in populations</u> *<u>with a risk factor and without</u>*, <u>to a risk factor value</u>** determined for the population impacted by this risk factor. The **Risk Factor** value here is 0 and more and defined as *(cases_with_risk - cases_without_risk) / cases_without_risk*. Here **Risk Causes / No Risk Causes** are numbers of causes (physiological parameters changes) determined for a disease in a population with a risk factor and without it using a formula presented above and are integer numbers (0,1,etc.). The formula is:

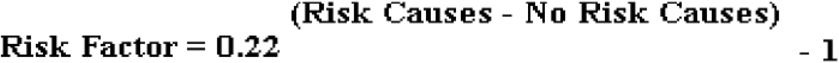

The formula is a foundation of the criteria to determine if the risk factor is really a disease cause. **<u>It connects physiological parameters changes in the human body to the risk factor which creates them</u>** for an individual. Using this formula researchers **can determine a number of physiological parameters changes the specific risk factor is causing in the human body**.

## Main Content

In this work, we will look at the causes of diseases in humans and will try to find a generic way to determine which possible reasons are causing a specific disease using mathematical statistics. The same rules derived in this study will likely apply to multi cell animals as well. We will build a model of non-infectious disease which, as every model, will be missing some details not considered important for the disease analysis.

1. Let us consider the human body as a physiological system placed inside an external environment that has the capability to impact the **physiological parameters** of the human body, called **parameters** further. This external environment, by acting on parameters of the human body, can cause a disease. If this environment does not negatively impact the human body, then the disease should not occur as there is no reason. In this model, we ignored the fact that even a healthy human body working in a non-harmful environment may have aging effects which cause diseases.
2. Let us describe the human body as a physiological system that has a set of certain parameters **An** as **{A1, A2, A3**,…, **An}**. Suppose the external environment causes a change in one and only one of these parameters, **Aj** where j E (1, n) and so others remain unchanged; then, this change should cause the same disease. In reality, we observe many different diseases, <u>so different diseases should be caused by changes at least in different parameters of</u> **<u>An {A1, A2, A3</u>**<u>,…,</u> **<u>An}</u>**<u>or by a group of some of parameters included in a set of</u> **<u>{A1, A2, A3</u>**<u>,…,</u> **<u>An}</u>**.
3. Suppose that each disease is caused by only one distinct parameter of the group **{A1, A2, A3**,…, **An}**. From experience, we know that the values of the parameters of the human body are usually random variables with a normal (Gaussian) distribution from Amin < Aj < Amax (for any j E (1, n)), such as weight and blood pressure variations, etc. Let’s recall that about 95.4% of all values of this parameter will be located within 2**σ** from a mathematical expectation **μ** of this parameter, 68.3% of values of this parameter will be located within 1**σ** interval from its a mathematical expectation **μ**. For a purpose of estimate, let’s accept that value of physiological parameter negatively affected by some external factor causing a disease is located beyond the 1**σ** interval meaning outside {-1σ, 1σ}
4. We take 1**σ** from a mathematical expectation **μ** as it is pretty far from average value of the parameter but is not exceeding the normal “healthy” range at the same time, also only a *small portion* (about 32%) of all values located in this interval. We use this range as also we consider a fact that *actions of external factors on human body is compensated by its capabilities to sustain physiological parameters* the same via homeostasis, for example a temperature of the body is around 98.6 F and can decrease or increase slightly but in a healthy person it tries to stay around this “normal” 98.6 F value. Homeostasis provides dynamic equilibrium for physiological parameters. The significant difference in the physiological parameters from their average “healthy” range under the actions of external factors is not likely due homeostasis.
5. Let us look at the case where the change in the parameter’s value **Ak** occurs in the positive direction and its value grows above 1**σ** and so will be located inside the interval where **Ak** E (**μ** + **σ, ∞**). The probability that parameters **Ak** will be in this zone is P(1< z) = Ф(z=3.99) - Ф(z=1) = 0.5 - 0.34 = **0.16** based on the normal distribution probability tables. The probability **0.16** that parameter **Ak** will have a value above **(μ** + **σ**) zone means that on average **one** or **two** persons **out of 10** should be sick with a disease if it is caused by a change *in one and only one* distinct parameter. The experience contradicts our original assumption that disease can be caused by just one parameter change, as for many *non-infectious* diseases, their probabilities in a society is about 5 per 100000 people (0.00005 for Leukemias), 303 per 100000 people (0.00303 for Multiple Sclerosis), or similar ranges and far not as high as 0.16. This contradiction implies that changes in **<u>one and only one distinctive physiological parameter is not enough to cause the majority of non-infectious diseases (where specific infections are not the main known cause) if the human body parameter’s change is just above the 1 σ (sigma) from math expectation μ in a positive(negative) direction</u>**. If we consider the possibility that the parameter can be lowered in the same fashion (to **μ** - **σ**) to the low border of its range, then the probability of these two events (**μ** + **σ** top and **μ** - **σ** bottom) would double and be 0.16 * 2 = 0.32, which is still very high compared to practical observations of *non-infectious* disease occurrences. Now, let’s us estimate what would happen if an physiological parameter after some negative action on it by external environment would be located beyond 2**σ** in a positive or negative direction? This would be practically outside of interval where *majority* of all possible values located. In this case P(2< z) = Ф(z=3.99) - Ф(z=2) = 0.5 - 0.477= **0.023**. That would mean that the non-infectious disease would be happening as often as 2 in one 100 people. This also contradicts to frequency of occurrence of most non-infectious diseases, for example it is about 3 per 1000 for Multiple Sclerosis (0.00303), etc. We can restate our conclusion that **<u>a change in</u> *<u>only one physiological parameter of human body by 1 σ (sigma) above math expectation cannot cause a majority non-infectious diseases as per observation of frequency of these diseases in a real life</u>***.
6. <u>We can state then that a disease generally is a result of change in multiple parameters of the group of</u> **<u>An {A1, A2, A3</u>**<u>,…,</u> **<u>An}</u>**. The probability of disease which is caused by multiple independent factors would be **P(D) = P(Ak1)** ^**×**^ **P(Ak2)** ^**×**^ **P(Ak3)** …^**×**^ **P(Akn)** where **kn** E (k1, n) is number of parameters of human body which were changed by more then 1 **σ** (sigma) in positive (or negative) direction. Based on the above calculations, we already know the probability of one parameter change above/below 1 **σ P(Awn) = 0.16** and let’s take it as an estimate that the same probability applies to each of the parameters. In this case, the probability of disease **P(D) = P(Awn) ^ n** (power of n), where **n** is the number of physiological parameters which change causes the disease.
7. From the above equation, we can obtain number **n** of physiological parameters of the human body (number of disease causes) which change during a disease as

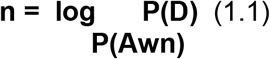

or 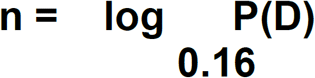, where P(D) is the frequency(rate) of the disease

The frequency(rate) of disease P(D) can be equal to 0.001 if it occurs in 1 in 1000 people. *<u>Number</u>* ***<u>n</u>*** *<u>of physiological parameters can be a real number and its needs to be approximated by closest integer numbers</u>* (for example 2.4 is approximated by 2, 2.7 is approximated by 3 as per usual math practices).

Now, P(Awn) = 0.16 was our theoretical estimate. Let’s try to determine the **P(Awn)** value more precisely from a real data observations. We know that occurrence of chronic diseases increases as people age and according to our model this means environment has time to change physiological parameters of the body during an individual’s live. Let’s do a practical estimate for **P(Awn)** based on children where less impact to physiological parameters is expected.

According to **NIH** (National Institute of Mental Health) occurrence of **autism** in boys is 36.5 per 1000 as year 2018 data, and in girls it is 8.8 per 1000 on the same year.

For boys 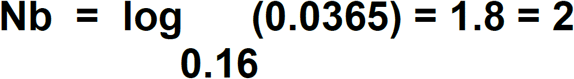 (2 physiological parameters causing **autism** in boys).

For girls 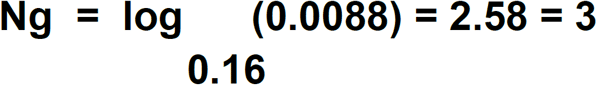 (3 parameters changes causing **autism** in girls)

If **Pb(D)** and **Pg(D)** is disease frequency for boys and girls accordingly then we know

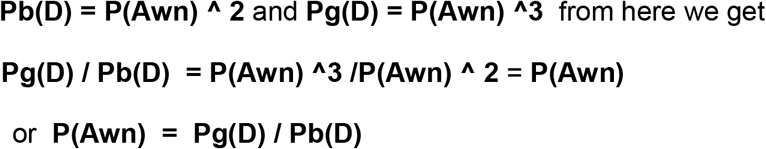

and substituting values for Pb(D) and Pg(D) we get

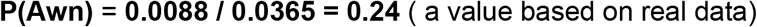

So based on the *data for a real disease* rate in humans (for autism) we get **P(Awn) = 0.24**. From practical usage perspective, to improve **n** value precision as an integer we reduce the value found to **P(Awn) = 0.22** which does not impact significantly the results. The final suggested formula for practical usage is following:

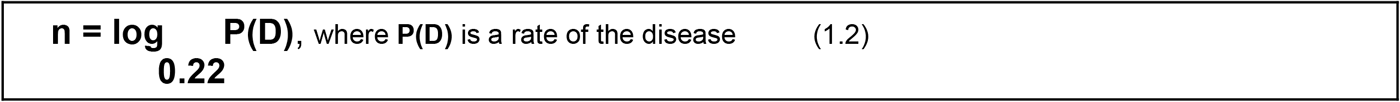

As already mentioned above, the frequency(rate) of disease P(D) can be equal to 0.001 if it occurs in 1 per 1000 people. *<u>Parameter n can be a real number and its needs to be approximated by closest integer numbers</u>* (for example if n = 2.4 is approximated by 2, n= 2.7 is approximated by 3 as per usual math practices).

We deduced a formula 1.2 based on our model. Based on this model the formula is expected to show **at least 2 physiological parameters changes** or more causing non-infectious diseases. Also, as we currently understand, the **same number of causes** is expected for each disease despite the fact that rate of the same disease **vary significantly**. Let’s see now how the formula 1.2 is matching the *real world data*:

### Coronary heart disease(CHD)

**Table 1.**

| Disease | Country / Region | Disease Rate<br>(number of sick<br>per 100 000<br>individuals) | Number of<br>disease causes<br>($\log_{0.22} P(D)$) | Comments |
| --- | --- | --- | --- | --- |
| Coronary heart disease(CHD) | Glasgow (UK) | 777 | $3.20 = 3$ | CHD is caused by 3 physiological parameters changes |
| same | Belfast (UK) | 695 | $3.28 = 3$ | CHD is caused by 3 physiological parameters changes |
| same | Barcelona (north eastern Spain) | 210 | $4.07 = 4$ | CHD is caused by 4 physiological parameters changes |
| same | Lille (northern France) | 298 | $3.84 = 4$ | CHD is caused by 4 physiological parameters changes<br>(French<br>Paradox) |
| same | Toulouse (south<br>western France) | **233** | 4.00 = **4** | CHD is caused<br>by **4**<br>physiological<br>parameters<br>changes<br>(French<br>Paradox) |

### Multiple Sclerosis

**Table 2.**

| Disease | Country /<br>Region | Disease Rate<br>(number of sick<br>per 100 000<br>individuals) | Number of<br>disease causes<br>(log P(D))<br>0.22 | Comments |
| --- | --- | --- | --- | --- |
| **Multiple**<br>**Sclerosis** | **Germany** | **303** | 3.83 = **4** | **4 physiological**<br>**parameters**<br>**changes in a**<br>**person causes**<br>**MS** |
| same | **USA** | **288** | 3.86 = **4** | **4 physiological**<br>**parameters**<br>**changes in a**<br>**person causes**<br>**MS** |
| same | **Canada** | **250** | 3.95 = **4** | **4 physiological**<br>**parameters**<br>**changes in a**<br>**person causes**<br>**MS** |
| same | **Switzerland** | **180** | 4.17 = **4** | **4 physiological**<br>**parameters**<br>**changes in a**<br>**person causes**<br>**MS** |
| same | **France** | **155** | 4.27 = **4** | **4 physiological**<br>**parameters**<br>**changes in a**<br>**person causes**<br>**MS** |
| same | **Netherlands** | **150** | 4.29 = **4** | **4 physiological**<br>**parameters**<br>**changes in a**<br>**person causes**<br>**MS** |

### Cancers

**Table 3.**

| Disease | Country /<br>Region | Disease Rate<br>(number of sick<br>per 100 000<br>individuals) | Number of<br>disease causes<br>(log P(D))<br>0.22 | Comments |
| --- | --- | --- | --- | --- |
| **Lung and Bronchus** | **New York, 2019** | **28.5** | **5.39 = 5** | **Lung and Bronchus Cancer is caused by 5 physiological parameters changes** |
| Lung and Bronchus | **Pennsylvania, 2019** | **35.7** | **5.24 = 5** | Lung and Bronchus Cancer is caused by **5** physiological parameters changes |
| **Female Breast** | **New York, 2019** | **18.2** | **5.69 = 6** | **Female Breast is caused by 6 physiological parameters changes** |
| Female Breast | **Pennsylvania, 2019** | **19.5** | **5.64 = 6** | Female Breast is caused by 6 physiological parameters changes |
| **Leukemias** | **New York, 2019** | **5.5** | **6.48 = 6** | **Leukemias is caused by 6 physiological parameters changes** |
| Leukemias | **Pennsylvania, 2019** | **6.3** | **6.38 = 6** | Leukemias is caused by 6 physiological parameters changes |

We can see from the calculated values in the tables that **Coronary heart disease(CHD) is caused by same number of 3 physiological parameters simultaneous changes** in UK but in France, and Spain the number of causes is actually higher as it **increased to 4** which explains so called *French Paradox* which is a fact that despite of high consumption of saturated fats, etc. the CHD occurrence in France (and as you can see in some places in Spain) is low comparing to other countries with high consumption of saturated fats as with UK in the above example. We can see that in France to get **CHD** people needs to experience a change of **4** parameters outside 1**σ** (sigma) where in UK only **3** parameters need to be affected and this means in UK some parameter is outside of 1**σ** (sigma) for most of people (level of “good” cholesterol possibly which in France is increased by high wine consumption ?).*The more physiological parameters changes are causing the disease the less the disease rate*.

We can see our deduced formula 1.2 and so our model gives results which are matching the predicted ones in the real world: 1) the *diseases are caused by multiple factors* (in the examples from **3 to 6**) and 2) Despite the differences in diseases occurrences the *number of physiological parameters changes (causes) is the same for the same diseases* for the same areas or located close (like New York and Pennsylvania States where **Lung and Bronchus cancer is caused by 5 physiological parameters changes** simultaneously occurring). 3) It can *explains why the occurrence of diseases significantly different* (2 to 3 times more) in different countries, for example like in UK and France for CHD, by differences in number of factors causing it. 4) It does not give the extremely high number of physiological parameters changes as it would be improbable to have them all happening at the same time (like 10 or 20 for example). **So practical data observations of non-infectious diseases look like matching the model we created and confirming it**.

## Experimental Prove of Formula for Disease Causes

The derived formula (1.2) **shows that the number of disease causes (physiological parameters changes) is related to the disease rate**.

Let’s prove that experimental data support the validity of the formula and a model it is based upon.

1. From a common sense, we can expect from this formula that **for different disease rates it would give same number of disease causes** - **Event A1**. If we take a look at Table 1 showing Multiple Sclerosis Rates and Causes we can see 6 countries having a different rate but all having 4 disease cause for MS. Let’s suppose the event **M** is that “a disease Rate is matching to the number of disease cause” is a coincidence. Let introduce the probability of this event **Pms** E {0,1} Let’s determine a probability **Pall** of the event that “for *all* countries disease rates for MS matches to the same number of disease causes N” which consist of independent events for each country.

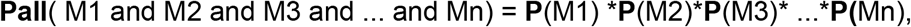

where M1, M2, M3, .. Mn are events that MS is matching to the same disease causes in different countries. Let’s take some value for **P(Mn)** from an interval {0,1} which is equal to **Pms** as 0.1(for a purpose of easy calculations) which is a pretty high probability the event is a coincidence and for an estimate let’s apply it to all **P**(Mn) where **n** = {1, n}. Then we get **Pall**(M1 and M2 and M3 and … and Mn) = **Pms**^**n** = 0.1^**n**. From the table 2 for MS diseases we can find already n = 6 at least so **Pms** = 0.1^6 and it means **Pall = 0.000001** which can be increased as we add more countries. It means the probability that events of type “MS disease Rate matching to the number of disease cause” happening together by coincidence is small **0.000001** and the complimentary event which is “event happens not by a coincidence” is 1 - **Pall** = 1 - 0.000001 ∼= 1 (it is very high so the fact is true) and this probability can be increased as we like with an addition of other countries causes. We can decrease P(Mn) from 0.1 to a smaller value and this will make **Pall** is even smaller. **<u>We proved that a match between the Disease Rates and a same number (</u>**<u>which we hope is a disease cause</u>**<u>) is not a coincidence but a fact</u>**. The probability of this fact P1 = 1
2. The formula calculating the **number of disease causes should give a positive number of diseases - Event A2**. We can see from Table for MS Disease Rates and Causes that it is the fact the number is positive. The probability of this fact P2 = 1 (means this fact is true, takes place)
3. The formula should give the **number of disease for a majority of disease as 2 and more** (as predicted by our model) - **Event A3**. We can see from Table for MS Disease Rates and Causes that it is the fact the number is more than 2. The probability of this fact P3 = 1
4. The **number of disease the formula gives should not give the amount exceeding 10s** - **Event A4** as this number of physiological changes will be unlikely to occur. We can see from Table for MS Disease Rates and Causes and other tables that it is the fact the number is less than 10. The probability of this fact P4 = 1
5. We don’t know how many causes is a valid number for a non-infectious disease (based on experience) but we can expect that **the difference in disease rates for same disease in women and men is caused by a small number of causes (physiological parameter changes) within a range {1, 3}** or so - **Event A5** as biochemically men and women are very similar but difference is in amount variations of small number of hormones (testosterone, estrogen, etc). We can see from autism causes above and calculations provided below for a Multiple Sclerosis disease’s causes that for a case of autism the number of causes (physiological parameters) differences between boys and girls is 1 cause, same for MS disease in men and women - the difference is in 1 cause only. The probability of this fact above P5 = about 1 (let’s say P2 ∼= 0.9) as we might need to see other similar cases. What is the probability that our formula is valid? It will be valid if probability of all events **A1, A2**,,,,, **An** above together is 1 and this probability is:

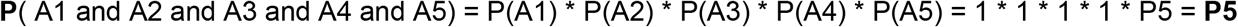

but **P5** is about 1 (or we take it as 0.9 or 0.95) so we can conclude that **experimental data support that our formula is valid with about 0.9-0.95 probability** or the chance that the derived formula matching the real world is about 90-95% which makes it predictions pretty precise for a practical usage (and the precision can be made higher with an increased number of data).

## Practical Conclusions from Disease Rate to Disease Cause Connection

We can make next conclusions out of analysis of the formula (1.2) above:

1. **<u>A specific disease only occurs if all of n found physiological parameters are changed above 1 sigma</u>** (are located outside of interval **(μ**-**σ**,**μ**+**σ**), slightly less than this). **<u>The disease cannot occur if only one 1 of n physiological parameters changed</u>**. *It resembles a lock with few wheels for unlocking* where each wheel is a separate physiological parameter. *Only unlocking ALL wheels of such a lock will cause a specific disease*. Based on this, disease cure (at least in the early stages) can occur if at least one of the parameters is within the normal range **(μ**-**σ**,**μ**+**σ**),. It also states that the human body has multiple levels of defense mechanisms, and the human body’s functions are supported at multiple levels which keeps a human body disease free moslty.
2. The **more** a rate of disease (occurrence per 1 thousand people, for example), the **less** the number of physiological parameters of the human body are changed to cause the disease. **<u>The more frequently a disease occurs the less a number of physiological parameters’ changes is causing it</u>**. This can be seen in Pic. 1 below, where **y** is the number of physiological parameters of the body and **x** is a rate disease.
3. The parameters found from the formula (1.2) are **physiological parameters** of the human body and not parameters of the external environment (it is not environmental factors, etc).
4. Physiological parameters needed to cause a disease are generally independent of each other (as they treated like independent events above).
5. In some cases, we can observe that in one area of some country, the population has a disease due to one number of physiological parameters changes and in another part of the country due to another number of physiological parameters changes. For example, in area 1 of the country, the disease occurs due to 4 physiological parameters changed and in area 2, the disease happen due 3 physiological parameters changed or the same can happen in the same area but at *different times* (e.g., 10 years ago or now). In this case, the additional physiological parameters (4th here) may be completely changed for the **entire a group of population** and only three remain unchanged. The simultaneous change in those 3 physiological parameters causes the disease.
6. Usually, diseases in practice occur due to changes in two to six physiological parameters changes in the human body depending on the disease rate. Infections are usually caused by a single factor and may occur very frequently.

**Pic. 1.**
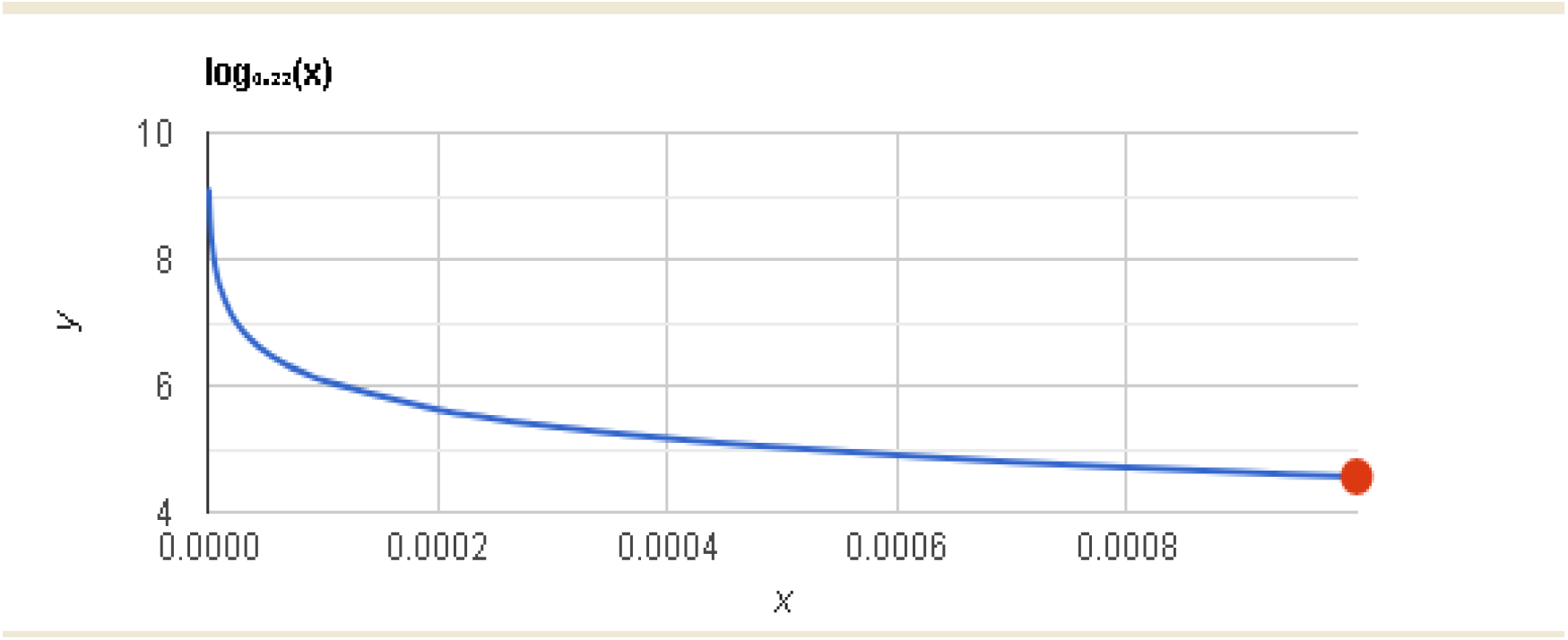
Number of disease causes (physiological parameters changed) related to a frequency of disease (disease rate). Here **y** (vertical) - number of causes, **x** (horizontal) - frequency of the disease (2 per 10000 as 0.0002)

Let’s see a practical application of the above formula (1.2).

In the USA, a rate of stroke in some of the states is 1.6-8 per 1000 people.

Let us determine the number of disease causes (physiological parameters changed) in the impacted people. Let us determine 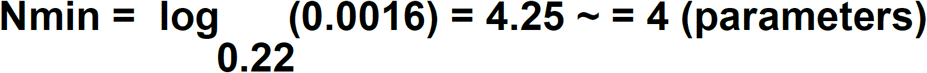

Let us determine 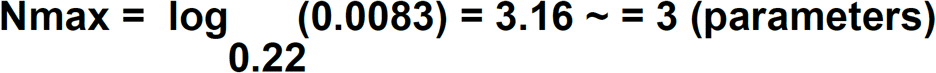

It means in some **US states a stroke is caused by 3 causes** and there are some where a stroke caused by 4 causes. The reason for this may be that in some states one physiological parameter out of 4 is already beyond 1-one sigma interval for a majority of population and does not protect it any more from stroke.

We need to notice here **that physiological parameters’ changes going beyond 1-sigma interval do not always mean that they are outside of usual “healthy” range** presented for a reference on different medical tests (blood work, urine test, etc). So in many cases **doctors will not notice these kind of changes**.

## Some Benefits of the Disease Causes Formula for Practical Applications

1. Based on the disease rate a derived formula (1.2) allows to determine a number of disease causes (as physiological parameters changes) in a specific population. **<u>It states that a rate of the disease is related to the number of physiological parameters changes which causing it and allows to find number of these physiological parameters changes</u>** and it means allows to find the number of disease causes.
2. As there is usually some research on the risk factors of certain diseases, the equation (1.2) above allows to use of these factors as if they are disease causes (physiological parameters changes) and prevent the disease from occurring in the population by keeping these physiological parameters in a healthy range {**μ**-**σ, μ**+**σ**}, slightly less. This reduces the significance of the specific risk factor in the analysis of disease causes, as there are *multiple* physiological parameters involved and causing the disease, but at the same time, **<u>it allows to take preventive measures for a disease by controlling these factors of risk to the point that disease never occurs</u>**. For example, if we know that stress hormones and blood pressure can increase the risk of stroke, then by controlling all these factors, we can increase the chance that the disease never occurs for a patient. Currently, we know that blood pressure is a risk factor for a stroke, *but we cannot say that it is a reason* for a stroke, as not all people with high blood pressure have strokes; if we decide *based on significance of research* that blood pressure is an important factor and can be one of the real physiological parameters the change of which causes the strokes, then controlling a blood pressure should prevent strokes from occurring. In addition, as a blood pressure is part of the set of other factors, the error in controlling it is compensated for by controlling other factors as well. Yes, we may have chosen a blood pressure as a causing disease factor wrong but we likely chose other factors correctly and controlling those factors as physiological parameters changes within 1-sigma interval will still prevent the disease.
3. **<u>The equation (1.2) above allows to use some of already found risk factors for a disease as disease causes as there are usually few (2 - 6) physiological parameters changes which are causing the disease and an error in choosing a risk factor as a disease cause gets reduced as if we chose some factors as physiological parameters correctly and some not but if we can control</u> *<u>all of them or most so they are within healthy range</u>***<u>{</u>**<u>μ</u>**<u>-</u>**<u>σ, μ</u>**<u>+</u>**<u>σ</u>**<u>}</u> **<u>then we still can prevent the disease even when one of physiological parameters chosen as a disease cause happens to be a wrong reason</u>**. It is usually sufficient to control only one right physiological parameter to prevent the disease, as mentioned above, as *all* needed to cause a disease.
4. This shows **<u>that majority of non-infectious diseases are caused by multiple causes (physiological parameters changes) which should take place at the same time</u>**. *It is not correct to search for just one specific cause* of the disease for a majority non-infectious diseases, which is currently done by multiple researchers.
5. This shows that many risk factors found for a specific disease do not work along and definitely, other factors exist. **<u>Controlling some of those risk factors can prevent disease from occurring though</u>**.
6. It shows that **<u>human body functions are provided by multiple mechanisms which back up each other</u>**.
7. **<u>As there are multiple physiological parameters’ changes needed to cause a disease it becomes easier to pick and choose them</u>** as they should be working as parts of the physiological mechanisms, and based on these physiological mechanisms, they can be selected more correctly as a set of multiple disease causes which should take place at the same time. Currently, we usually select a factor as a potential cause for a disease based on studying the factor as a standalone.
8. As a disease is caused by physiological parameters changes beyond **1-sigma** interval the **reference ranges on medical tests (blood work, urine**,**etc.) may need to be updated** so doctors can see when a specific parameter goes beyond this range and are able to notify patients that they are moving to some disease and need to take corrective actions.

## Formula for Determining if a specific Risk factor is causing a Disease

Let’s deduce the formula connecting a risk value with an associated change in few out of **n** physiological parameters of the human body. Let’s take 100 people(or 100%) *without* a risk factor and let the number **m** (or **m%)** be a number of sick people having a certain disease (this is usually found through a medical research). Let’s take another 100 people(or 100%) *with* a risk factor and let the number **l** (or **l%)** be a number of sick people having the same disease. Let’s look at the cases where where **l > m (**which is usually practically occurring as we estimate how the specific risk increases number of sick people).Then the risk **K** is:

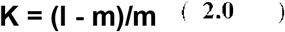

For the those not having a risk factor a probability of disease then **Pnr(D) = m/100** and with a risk factor **Pr(D) = l/100**

From our disease factor formula we know that

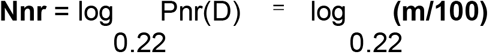

from here we get **m** = 0.22^Nnr^ · 100

Same way we get for Nr (number of factors causing disease with a risk factor present)

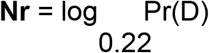

from here we get **l** = 0.22^Nr^ · 100

Substituting the **l** and **m** values into formula 2.0 above we get:

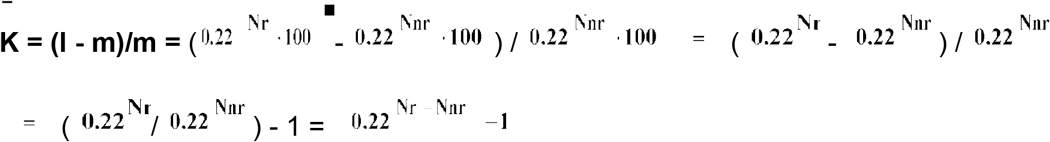

We get a **<u>formula (2.1) connecting a number of physiological parameters causing a disease with a risk factor K creating these physiological parameters changes</u>** where **Nr** is number of physiological parameters causing disease *with a risk factor* present and **Nnr** is a number of physiological parameters causing disease *without a risk* factor present.

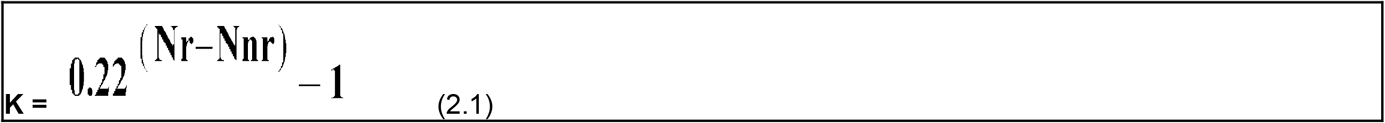

Where **Nr > Nnr, Nr** and **Nnr** E {1,2,3,4,5,..}

**<u>The formula 2.1 shows that with an increase of disease risk factor K, the number of physiological parameters Nr necessary to cause the disease gets decremented by only the integer numbers, starting with 1. The disease risk factor’s K value associated with each decrease in number of physiological parameters Nr, takes up the</u> *<u>one and only possible theoretical value which uniquely matching to every integer number by which the physiological parameters amount gets reduced</u>***. For example, as it will be shown below, for a risk factor caused by 1 physiological parameter decrease (in other words its disappearance) the theoretical value of disease risk **K = 3.55** (or 355%).

## Criteria to Determine If a Risk Factor is a Disease Cause

If we are doing an experiment and found from it a number **Nr** of physiological parameters causing a disease in a population *with* risk factor and we found a number **Nnr** of physiological parameters causing the same disease *without* a risk factor and now we want to check if the risk factor we working with impacts only **one** physiological parameter of the body, then **Nr - Nnr = -1** (−1 as the **Nr** should be less than **Nnr** because with an increased risk number of physiological parameters changes gets reduced as per disease frequency formula (1.2), also a number of parameters are integers only 1, 2, etc). Then for n = 1 we can write down

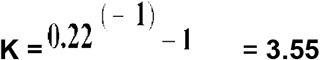

This means **<u>if a risk factor is causing only one physiological parameter change in the human body and is really causing a disease then the risk factor theoretical value should be 3.55 (or 355%)</u>**. We know that a risk factor is causing a disease if the factor is an external one, as external factors are causing a change of physiological parameters and this way are causing a disease as per our model. For example, a solar radiation is an external factor which may cause a skin cancer and the criteria above allows to determine if this radiation is a cause of the disease or not.

Then for n = 2 we can write down

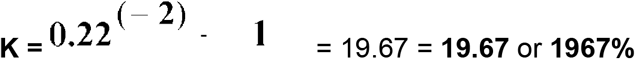

This means **<u>if a risk factor changing 2 physiological parameters is causing a disease then its value should be theoretically equal 19.67 (or 1967%</u>)**

The author suggests to **use a rule that the practical risk factor can be located within a theoretical +/-50% range** as it is clear that practical data will be somewhat different than theoretical. For example, for a risk factor responsible for just one physiological parameter change its practical value should be within range {355 -178, 355 + 178} or between **177% and 533%**

## Smoking is one cause of lung cancer

Let’s look at one example of estimation if a risk factor is really causing a disease.

For people *who smoke the risk of lung cancer* is at least **15** times bigger than for non-smoking people. So if number of non-smoking sick people is **m** then **l = 15 *m** and risk **K = (15m - m) / m = 14 or 1400%** it is clear it does not match to 1 physiological parameter change which is 355% but pretty close to 1967% for 2 physiological parameter changes. So let’s estimate the deviation as **(1967 - 1400) / 1967 = 0.29 or 29%** which is less than 50%

So **<u>the estimate shows that the smoking is really one of the causes of lung cancer</u>** and impacting **2** disease causing physiological parameters.

## Low beta-carotene is one cause of breast cancer

Let’s look at another example of risk factor estimation. The research shows that women having beta-carotine in lower quartile of the range are more than **220%** likely to get a *breast cancer* than those having *beta-carotene* in the top quartile. It means the value is pretty close to 355% for 1 physiological parameter change and is really within 50% of the practical range as it is above 177% and less then 533% for 1 physiological parameter change as shown above. Based on this risk factor estimate we conclude **<u>that low beta-carotene is one of multiple causes of breast cancer</u>**.

Accordingly to early made conclusions **if women control *beta-carotene* level so it is pretty high (within one sigma) then breast cancer should not happen unless there is a damaging factor** (like a carcinogen, etc) impacting them and in this situation it is necessary to control 2 or more physiological parameters causing the disease in order not to get sick and remove action of carcinogen. This will be discussed further.

## Examples of practical applications for the formula of number of disease causes

Let’s us see some examples of practical applications for the number of disease causes formula (1.2) derived in this work.

### Multiple Sclerosis

In USA this disease happen in 277 people per 100000, in women the disease happens 3 times more often than in men

Let’s calculate the number of physiological parameters which changes beyond 1-sigma interval are causing the disease in men and women:

If we consider as x - number of men sick, then women are 3x and then out of equation x + 3x = 277 we get 69.25 = 69 men per 100000 people sick and for women 3x = 3 * 69.25 = 207.75 women per 100000 people sick

Number of factors causing **Mutliple Sclerosis in men then 5 :**

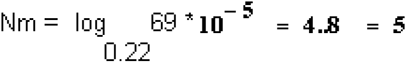

Number of factors causing **Mutliple Sclerosis in women then 4 :**

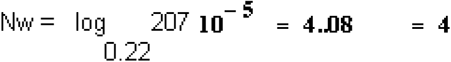

In women Multiple Sclerosis happens *more often* because to cause it the *one less* physiological parameter change outside of 1 sigma is required. **It requires 4 changes to physiological parameters for women to get sick vs. 5 physiological parameters changes for men to get sick with the Multiple Sclerosis**.

It mean the 5th physiological parameter for every women either always outside of 1 sigma during the disease vulnerability times or possibly does not exist at all.

### Alzheimer

In USA this disease happen to women in 108 people per 10000, in men 56 per 10000 people. Doing calculation we get:

For women **Alzheimer happens because of 3 internal parameters changes**

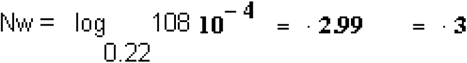

For men **Alzheimer happens because of 3 internal parameters changes**

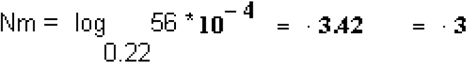

**Alzheimer disease happens to men and women due to 3 physiological parameters changes** beyond 1-sigma interval. Alzheimer **is not happening just because one protein accumulation** in the brain according to this.

### Atrial fibrillation

In USA this disease happen to people under age 65 as 1 in 50 people. Over age of 65 this disease happen to 1 in 10 people.

Doing calculation we get:

For age **under 65 Artrial fibrilation happens because of 3** parameters changes

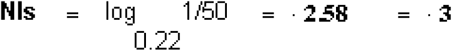

For people **over 65 Artrial fibrilation happens because of 2** parameters changes

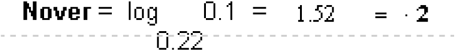

With aging the environmental factors(as per our model) reduce a number of physiological parameters of the body which needs to be changed outside of 1-sigma of their measurements in order to cause the disease from 3 physiological parameters changes to 2 physiological parameters changes and **this explains why atrial fibrillation happens more frequently for people over 65** years old as the less number of physiological parameters changes required to cause a disease then more a rate of the disease.

## Three causes of Atrial fibrillation

There are multiple risk factors found in medical research which increase a disease rate of atrial fibrillation(**AF**) in person but not all them matching criteria set by formula 2.1 regarding a risk factor value which required for the risk factor to be a cause of the disease. Let’s see some risk factors of atrial fibrillation which actually do pass this criteria. There should be 3 physiological parameters changes they impact as we found above, to cause the AF disease.

People with Obstruction Sleep Apney (OSA) found 5 times more likely to develop AF than without OSA. If **x** - number of people without OSA then K = (5x/100 - x/100) / (x/100) = 4 or **K% = 400%** which is in range between **177%** and **533%** and per discovered above criteria for 1 physiological parameter risk **this means that Obstruction Sleep Apney is one of reasons of atrial fibrillation**. We can count it as reason 1 of disease.

It was found that epicardial fat increase over 1-standard deviation was associated with 5.4 fold high odds of persistent AF. Calculating in same way as above K = ((5.4x - x) / 100 : x/100) = 4.4 or **K% = 440%** which is in range between **177%** and **533%**.

**It means an epicardial fat increase over 1-standard deviation is causing atrial fibrillation**. We can count it as reason 2 of the AF disease. The fact that *the fat should be increased above 1 sigma to cause the disease also is according to our model and formula 1*.*2*

Aldosterone found as risk factor for AF as it has been reported to create 12 fold higher risk of AF in hypertension patients with primary hyperaldosteronism when compared with patients with essential hypertension. Calculating K = (12x - x)/100 / x/ 100 = 11 or **K% = 1100**. As we can see the risk factor is too high for 1 physiological parameter change, but looks like within a range for 2 physiological parameters changes. Let’s estimate a deviation from a theoretical risk value of **1967%. dK = (1967 - 1100) / 1967 = 0.44** or **44%**, as it is below 50% we can consider the risk in the range for 2 physiological parameters changes as it was shown above. It means **Aldosterone is a one of the causes for atrial fibrillation**. We can count as reason number 3.

According to presented in this article model the *3 risk factors* (or here 3 matching them physiological parameters changes) *should exist at the same time to cause the disease* and this explains why they are called just risk factors as if one of them is present does not mean other 2 exist at the same time and so person should not be sick with AF. It means the **atrial fibrillation develops in people if they have Obstruction Sleep Apney(OSA) *AND* high level of aldosterone *AND* and an epicardial fat increase over 1-standard deviation**. Or they can have high level of aldosterone and one additional factor from above as aldosterone covers 2 physiological parameters changes of parameters as to cause the AF disease we need 3 physiological parameter changes for people under 65.

Now, we can see above that for people over 65 the AF requires 2 physiological parameters changes only and this means that these people should have one of the discovered causes applicable to this entire group. Which causation factor(physiological parameter) is changed for entire group needs to be found. It looks like a *good hypothesis* to research is that *an epicardial fat increase over 1-standard deviation is applicable to majority of people over 65* and causing an increase in frequency of atrial fibrillation in this group of population as OSA probably does not apply to all people after 65, and it is doubtful that most people over 65 have high level of aldosterone levels as well.

### Coronary Artery Disease (CAD)

In USA this disease happen to 20.1 **Million** of adults over age 20. It is approximately in 7.2% of adult population in USA.

Doing calculation we get that **Coronary Artery Disease happens due to 2 causes** (2 physiological parameters changes beyond 1-sigma interval)

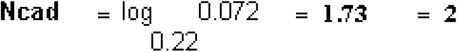

Only 2 physiological parameters parameters changes outside of 1 sigma *needed at the same time* to cause the CAD disease and **it explains why CAD disease so frequently occurs** in population as the less the number of causes for a disease the higher rate of the disease in population.

As we can see in few example above some physiological parameters are changed for the **entire group** of particular population - men/women, older 65/under 65. etc. and **<u>this causes different disease rates in these groups of population</u>**.

## Conclusions

This work showed that non-infectious diseases are caused by multiple simultaneously taking place factors and this factors are the physiological parameters changes approximately beyond 1 sigma interval. It explains why some disease occurring in men or women, different world regions, etc. at different rates by showing that in many cases it is due to different number of factors which cause a disease and the work allows to determine these specific differences in numbers. The model introduced here allows to improve efficiency of medical research by allowing to convert disease frequency information into a useful information about a physiological mechanism structure of a specific disease first and then allows to find disease causes using provided criterias by applying them to the results of existing or new medical researches. This work is covering some gap in understanding of disease causation which is a significant roadblock of modern medical research. By introducing a concept of multiple factors causation it allows to make an easier selection of the risk factors as diseases causes as it shows that there is a room for an error in this selection and this determination of causes should allow more effective disease prevention and potentially their cure at early stages by controlling appropriate factors.

Alan Olan – 2019

## Data Availability

All data produced in the present work are contained in the manuscript

https://www.worldlifeexpectancy.com/france-coronary-heart-disease

## Ethical Compliance

All procedures performed in studies involving human participants were in accordance with the ethical standards of the institutional and/or national research committee and with the 1964 Helsinki Declaration and its later amendments or comparable ethical standards.

## Author Contributions

Alan Olan is the sole contributor to this study

## Disclaimer

This work is not intended to be used in diagnosing and treatment of any disease in any patient, those actions should be done by qualified professionals only. The work is intended as a reference for the researchers only. The author is not responsible for any issues created by unintentional usage or consequences of this work.

